# Kinetics and performance of the Abbott Architect SARS-CoV-2 IgG antibody assay

**DOI:** 10.1101/2020.07.03.20145722

**Authors:** Fergus Hamilton, Peter Muir, Marie Attwood, Alan Noel, Barry Vipond, Richard Hopes, Ed Moran, Nick Maskell, Deborah Warwick, Mahableshwar Albur, Jonathan Turner, Alasdair MacGowan, David Arnold

## Abstract

**Objectives:** To assess the performance (sensitivity and specificity) of the Abbott Architect SARS-CoV-2 IgG antibody assay across three clinical settings.

**Methods:** Antibody testing was performed on three clinical cohorts of COVID-19 disease: hospitalised patients with PCR confirmation, hospitalized patients with a clinical diagnosis but negative PCR, and symptomatic healthcare workers (HCW’s). Pre-pandemic respiratory infection sera were tested as negative controls. The sensitivity of the assay was calculated at different time points (<5 days, 5-9 days, 10-14 days, 15-19 days, >20 days, >42 days), and compared between cohorts.

**Results:** Performance of the Abbot Architect SARS-CoV-2 assay varied significantly between cohorts. For PCR confirmed hospitalised patients (n = 114), early sensitivity was low: <5 days: 44.4% (95%CI: 18.9%-73.3%), 5-9 days: 32.6% (95%CI, 20.5%-47.5%), 10-14 days: 65.2% (95% CI 44.9%-81.2%), 15-20 days: 66.7% (95% CI: 39.1%-86.2%) but by day 20, sensitivity was 100% (95%CI, 86.2-100%).

In contrast, 17 out of 114 symptomatic healthcare workers tested at >20 days had negative results, generating a sensitivity of 85.1% (95%CI, 77.4% - 90.5%). All pre-pandemic sera were negative, a specificity of 100%. Seroconversion rates were similar for PCR positive and PCR negative hospitalised cases.

**Conclusions:** The sensitivity of the Abbot Architect SARS-CoV-2 IgG assay increases over time, with sensitivity not peaking until 20 days post symptoms. Performance varied markedly by setting, with sensitivity significantly worse in symptomatic healthcare workers than in the hospitalised cohort. Clinicians, policymakers, and patients should be aware of the reduced sensitivity in this setting.

## Introduction

The accuracy of antibody tests for SARS-CoV-2 is a matter of huge public health importance. Although many cohorts have reported rapid IgG responses to infection with in-house, commercial, or lateral flow assays, there has been little external validation of the commercial assays.^1–4^ Public Health England approved the Abbott SARS-CoV-2 IgG antibody test based on testing 96 COVID-19 patient samples and 760 presumed negative samples. They found a sensitivity of 93.9% (95%CI 86.3-98.0), and specificity of 100.00% (95% CI 95.9-100.0) by 14 days post symptom onset.^5^ This antibody test can run on the commercial Architect platform, allowing for rapid rollout and testing. However, there has been limited external evaluation of this assay. In this paper, we report the kinetics and performance of this assay in three populations: confirmed (PCR +ve) and suspected COVID-19 patients, confirmed (PCR +ve) healthcare workers, and pre-pandemic controls with respiratory infection.

## Methods

### Study design

This study aimed to estimate the kinetics of the antibody response (i.e time taken to seroconvert) with the Abbott Architect SARS-CoV-2 IgG antibody assay, and estimate the sensitivity of this assay.

This study is reported in line with the StarD guidance on diagnostic accuracy studies (Appendix 1)

### Sample provenance

Samples came from four groups. The first two groups consisted of 94 patients with PCR-confirmed COVID-19 and 35 patients who were clinically suspected to have COVID-19 but tested PCR negative. These two groups were recruited prospectively in to the DISCOVER study at North Bristol NHS Trust for which HRA Approval was granted by the South Yorkshire Research Ethics Committee (20/YH/0121). Clinical and demographic features were recorded daily during in-patient stay and subsequently at out-patient follow-up clinics. Samples were submitted for PCR testing at admission or first presentation. Further samples were tested where initial samples tested negative for SARS-CoV-2 RNA. Serum or plasma samples were collected at various time points during in-patient and out-patient follow-up (Appendix 1)

The third group consisted of 114 healthcare workers at North Bristol NHS Trust who tested SARS-CoV-2 PCR positive during investigation of suspected COVID-19, and from whom serum or plasma samples were tested for SARS-CoV-2 antibody as part of NHS England’s healthcare workers testing programme. All antibody tests in this group were performed at >20 days after PCR testing.

Due to data protection concerns, we could not access their hospitalization records, but to our knowledge only a very limited number of HCWs were hospitalised with COVID-19, the vast majority having mild disease.

The fourth group consisted of 20 a longstanding prospectively collected bank of clinical samples from patients presenting with parapneumonic effusions/pleural empyema to the Academic Respiratory Unit, University of Bristol prior to the COVID-19 pandemic.

### Statistical approach

The sensitivity of the Abbott SARS-CoV-2 IgG assay was estimated with 95% Confidence Intervals at different time points post symptom onset (DISCOVER patients) or first PCR positive result (healthcare workers). As all healthcare workers tests were more than 20 days after the PCR date, we assumed the onset date for all healthcare workers was more than 20 days.

Specificity was estimated using the pre-pandemic controls with parapneumonic effusions/pleural empyema. For calculation of sensitivity, only PCR confirmed DISCOVER patients were included. Comparisons were made between confirmed and suspected cases, and relevant comorbidities in the hospital cohort (severity of disease, age, gender) using the log rank test. All analysis was performed in R version 4.00 (R Foundation for Statistical Software, Vienna), using the packages “tidyverse”, “survival”, and “bootLR”. Confidence intervals around point estimates were generated using bootstrapping.

### SARS-CoV-2 antibody testing

The Abbott SARS-CoV-2 IgG assay was performed on the Abbott Architect analyser, according to the manufacturers instructions. The assay is a chemiluminescent microparticle immunoassay for detection of IgG in serum or plasma against the SARS-CoV-2 nucleoprotein. For the healthcare worker samples, fresh plasma was used; for the other two groups, plasma stored at - 20C was used.

### SARS-CoV-2 RNA PCR

Total nucleic acids were recovered from nose and throat swabs in viral transport medium or lower respiratory samples using a KingFisher Flex (Thermofisher) and tested for SARS CoV-2 RNA by real time PCR. During the period of this study SARS-CoV-2 RNA detection was performed using a RdRp real-time PCR ^6^ or the RealStar SARS CoV-2 real-time PCR (Altona Diagnostics) which detected dual E gene and S gene target sequences.

Specificity was estimated using the pre-pandemic controls with parapneumonic effusions/pleural empyema. For calculation of sensitivity, only PCR confirmed DISCOVER patients were included, to ensure a robust ‘true positive’. Comparisons were made between confirmed and suspected cases, and relevant comorbidities in the hospital cohort (severity of disease, age, gender) using the log rank test.

## Results

In total, 263 individual tests were performed, on 241 individuals. Assay sensitivity is shown in Table 1 for the three separate cohorts. There was a marked difference in performance between hospitalised patients and healthcare workers. For confirmed PCR+ cases, all antibody tests performed at >20 days were positive, whereas for healthcare workers 17 out of 114 tests performed were negative.

**Table 1:** Summary of all results

| Cohort: | PCR+ hospitalised patients<br>(n = 114) |  | PCR- hospitalised patients<br>(n = 35) |  | Healthcare worker testing<br>(n = 114) |  |
| --- | --- | --- | --- | --- | --- | --- |
| Days from onset: | IgG+/total tested | Sensitivity (CI's) | IgG+/total tested | Sensitivity (CI's) | IgG+/total tested | Sensitivity (CI's) |
| <5 | 5/10 | 44.4%<br>(18.9%-73.3%) | 1/8 | 12.5%<br>(2.2% - 47.1%) | n/a | n/a |
| 5-9 | 14/43 | 32.6%<br>(20.5%-47.5%) | 4/14 | 28.6%<br>(11.7% - 54.6%) | n/a | n/a |
| 10-14 | 15/23 | 65.2%<br>(44.9%-81.2%) | 4/5 | 80%<br>(37.6%-96.4%) | n/a | n/a |
| 15-20 | 8/12 | 66.7%<br>(39.1%-86.2%) | 1/2 | 50%<br>(9.5%-90.5%) | n/a | n/a |
| >20 | 26/26 | 100%<br>(86.2%-100%) | 5/6 | 83.3%<br>(43.6%-97.0%) | 97/114 | 85.1%<br>(77.4% - 90.5%). |
| >42 | 24/24 | 100%<br>(87.1%-100%) | 5/6 | 83.3%<br>(43.6%-97.0%) | 55/66 | 83.3%<br>(72.6% - 90.4%) |

### DISCOVER cohort

166 participants enrolled in the DISCOVER, cohort, of which 123 had the assay performed during their hospital stay, with 5 subsequently tested for the first time at a follow up clinic more than four weeks after admission. Age, gender, and comorbidities are described in Table 2. The cohort had a median age of 58, and comorbidities were common. 13 patients (7.9%) went to intensive care, while 15 patients died.

**Table 2:** Demographics of the DISCOVER cohort

| Characteristic | PCR positive , N = 125 | PCR negative, clinically suspected, N = 41 |
| --- | --- | --- |
| Age (18+) | 61 (47, 76) | 53 (41, 64) |
| Sex |  |  |
| Male | 72 (58%) | 20 (49%) |
| Female | 53 (42%) | 21 (51%) |
| Outcome at 28 days |  |  |
| Alive | 107 (87%) | 41 (100%) |
| Dead | 16 (13%) | 0 (0%) |
| Ongoing follow up | 2 | 0 |
| Level of care |  |  |
| Ward based | 110 (89%) | 35 (85%) |
| Intensive care | 9 (7.3%) | 4 (9.8%) |
| High Dependency | 4 (3.3%) | 2 (4.9%) |
| Ongoing follow up | 2 | 0 |
| Diabetes status: |  |  |
| None | 102 (82%) | 29 (71%) |
| T1DM | 2 (1.6%) | 1 (2.4%) |
| T2DM | 21 (17%) | 11 (27%) |
| Heart disease: | 37 (30%) | 6 (15%) |
| Chronic Lung disease: | 37 (30%) | 7 (17%) |
| Severe Liver disease: | 4 (3.2%) | 1 (2.4%) |
| Severe kidney impairment (eGFR< 30 or dialysis) | 14 (11%) | 4 (9.8%) |
| Hypertension: | 34 (27%) | 11 (27%) |
| HIV positivity: | 2 (1.6%) | 0 (0%) |
| <sup>†</sup> Statistics presented: median (IQR); n (%) |  |  |

Early sensitivity was low, but all samples collected >20 days post symptom onset were positive. Two of these later tests were performed as inpatients, with the remaining 25 performed in a follow up clinic. Table 3 provides the summary of the DISCOVER cohort sensitivity by time of onset. After 20 days, the calculated sensitivity was 100% (87.5% - 100%).

**Table 3:** Validation of the Abbot Architect assay (as of 18th of June 2020)

| Authors | Positive samples | Negative samples | Results: |
| --- | --- | --- | --- |
| Public Health England,<sup>5</sup> | 122 samples from 31 PCR-confirmed and symptomatic patients. | 1070 samples, (997 pre pandemic) | Specificity 99.6%, Sensitivity 86.6% day 8-13, 100% > day 14 |
| Paiva et al<sup>13</sup> | 113 samples from 73 PCR-confirmed hospitalised cases | 1,182 samples (1,063 pre-pandemic) | Specificity 99.62%, Sensitivity 44.8% day 7, 100% >day 15 (28 samples >15 days) |
| Perkmann et al<sup>14</sup> | 65 PCR+ or symptomatic donors/patients (mostly non-severe) | 1,154 samples, all pre-pandemic | Specificity 99.2%, Sensitivity 84.6% (all samples > 14 days) |
| Chew et al<sup>15</sup> | 117 symptomatic patients (admitted) | 163 non-COVID patients | Specificity 100%, Sensitivity 43.6% 7-13 days, 84.2% >14 days. |
| Bryan et al<sup>16</sup> | 685 samples from 125 PCR positive patients. | 1,020 samples, all pre-pandemic | Specificity 99.9%, 100% sensitivity at day 17. |

Cumulative seroconversion for the PCR positive cohort is shown in Figure 2. The median date of seroconversion was 13 days (IQR 12-15).

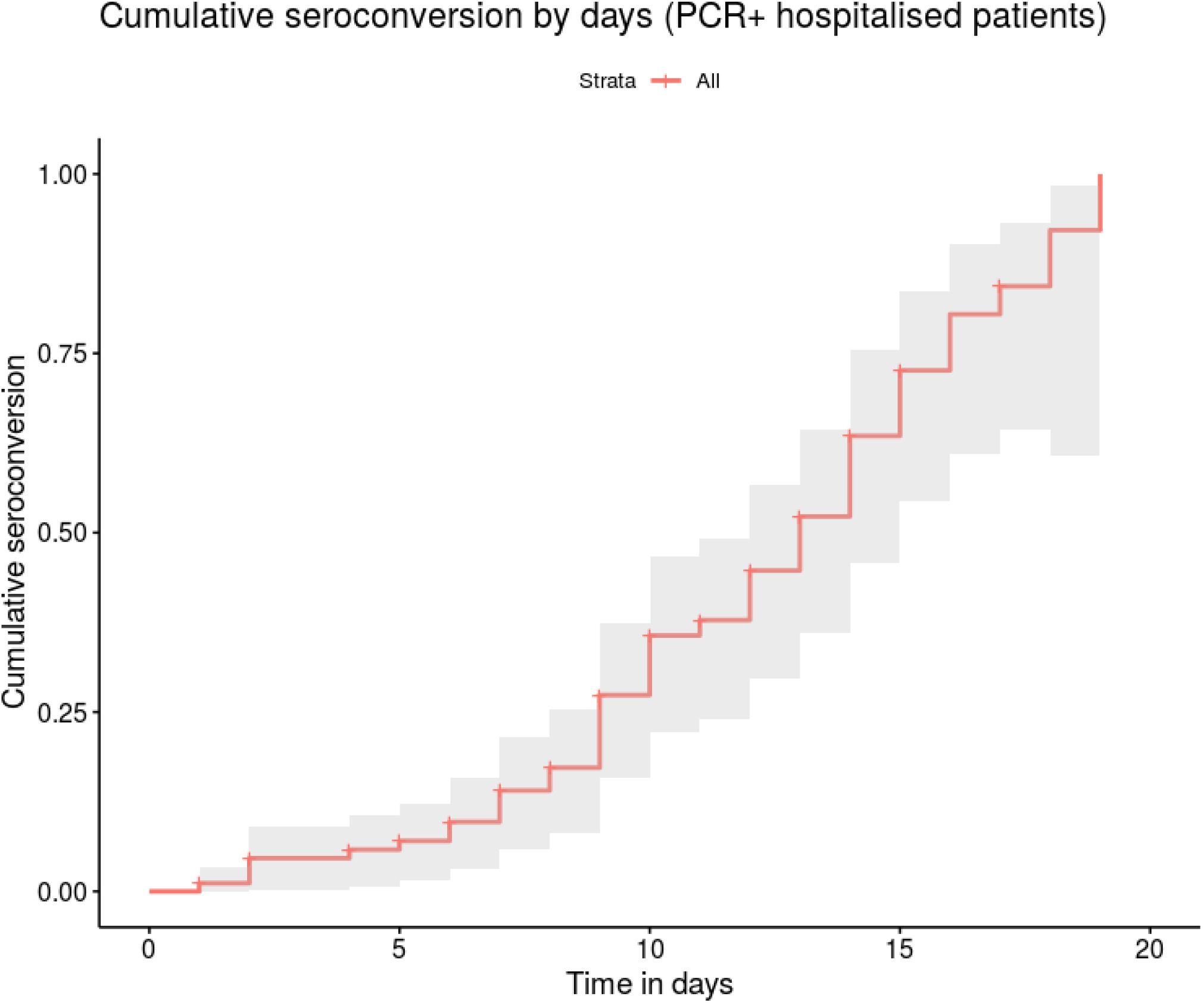

### Suspected vs proven disease

Seroconversion rates were similar between clinically suspected and PCR proven disease. Figure 2 shows the cumulative seroconversion, which was not statistically different between proven and suspected cases.

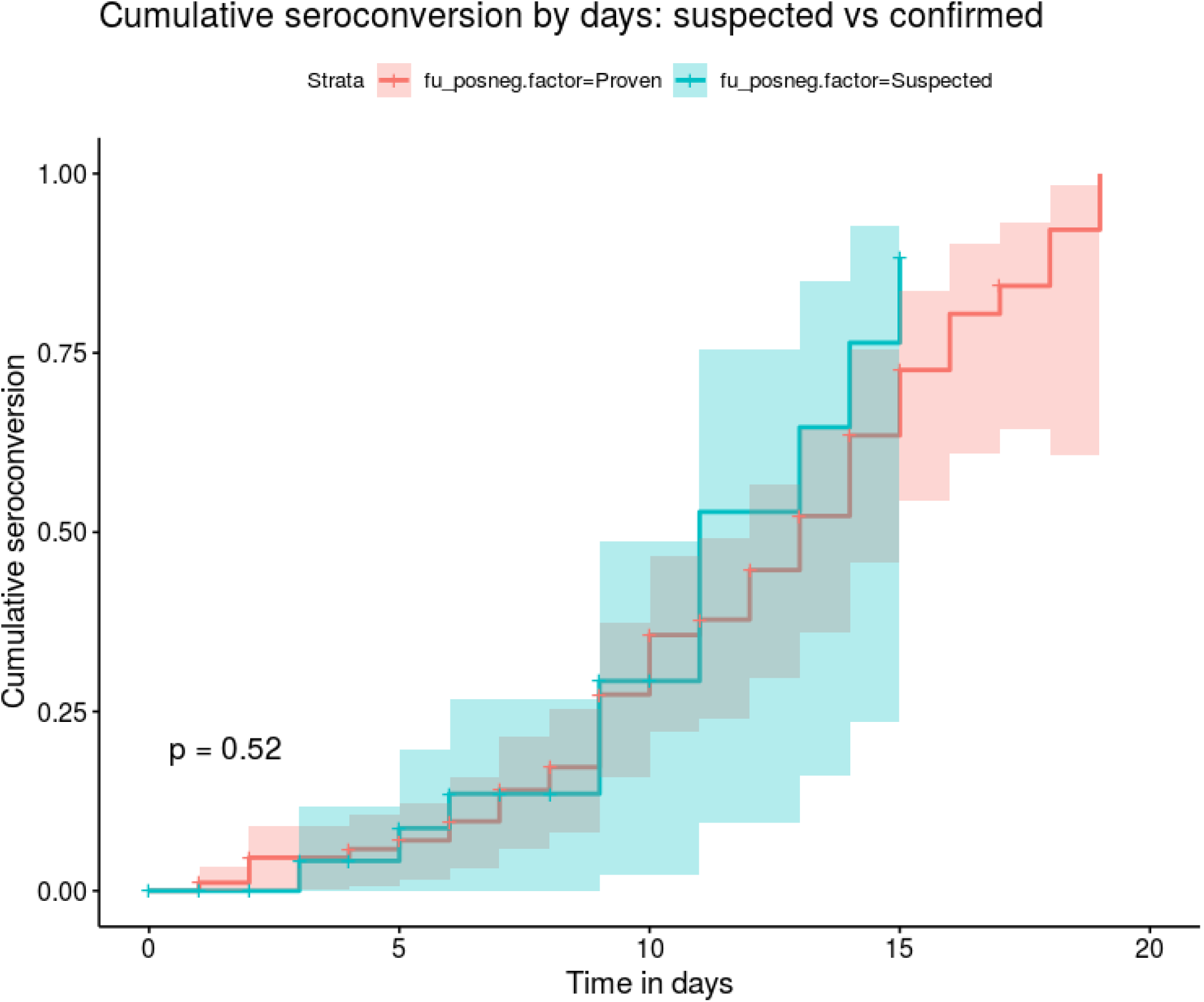

### Comorbidity

There was no difference in seroconversion between those who died (p = 0.97), went to intensive care (p = 0.40), gender (p = 0.42), or age (>65 years old, p = 0.42).

### Healthcare worker testing

In the healthcare workers testing cohort, 114 healthcare workers had positive PCR testing and subsequently went on to have antibody tests. The median time to test was 45 days, and all negative antibody tests were more than 32 days after the PCR result, but under 60 days. 97 of these antibody tests were positive, with 17 being negative. The sensitivity of the assay in this context was 85.1% (77.4% - 90.5%).

### Historic respiratory infection controls

All 20 pre-pandemic sera from the Pleural Investigation Database were negative. This corresponds to a specificity of 100% (83.9% - 100%).

## Discussion

Our results describe real world performance of the Abbott Architect SARS-CoV-2 IgG assay. There was a significant difference in timing and overall rate of seroconversion between healthcare workers, who had predominantly mild disease and hospitalised cases, with all hospitalised patients tested after 20 days having a positive test, but only 83% of symptomatic healthcare workers having a positive result at this point. Interestingly, seroconversion dynamics seemed similar in PCR negative and PCR positive cases, suggesting clinical diagnosis is accurate for COVID-19.

### Comparison with previous literature

There is limited literature on the performance of this and other antibody assays, and the kinetics of seroconversion to SARS-CoV-2 more generally. Many papers suggest IgG is detectable in around 95% of cases by around day 14^1,2,8–10^ but these analyses are based predominantly on hospitalised patients.

However, some papers suggest a slower IgG response, with a cohort of patients who never seroconvert. It is not known if these discrepancies are due to differential assay or differences in the patient populations studied.^1,11^. For the Abbott SARS-CoV-2 IgG assay Public Health England performed internal validation on 96 patients samples and 760 convalescent samples and found a sensitivity of 93.4% (95%CI 85.3 - 97.8), and specificity of 100.00% (95%CI 99.1-100.0). At the time of writing, four other groups have published data on this assay, although none in a UK population. Their data is summarised in Table 3. All groups found specificity was excellent, although the sensitivity of the assay varied widely. Unfortunately, most data included was from hospitalised patients, with only one paper definitively including 46 non-hospitalised patients^14^, with the sensitivity in that paper being similar to ours (84.6%, 95% 73.6% - 92.4%). This study thus augments the available data on assay sensitivity in different patient groups. Indeed our study reports the largest cohort thus far, with 203 PCR confirmed individuals, from both mild and more severe cases.

One explanation of reduced assay sensitivity in HCWs is simply that some milder cases simply do not produce significant IgG, which has been reported by other UK studies^11^though not others.^9,10^. Intriguing recent findings have suggested that some contacts of patients with COVID-19 who fail to seroconvert have evidence of T cell response to COVID-19 already, perhaps suggesting prior T cell immunity due to previous infection with other human coronaviruses or development of non-humoral immunity^12^.

### Strengths and weaknesses

The major strength of our cohort is the size, and the inclusion of milder patients that have been absent in many previous studies, and are a recommendation of a recent Cochrane review.^7^ As the clinical use of antibody testing is likely to be in patients without hospitalisation, this data is invaluable. All previous papers except one have had limited clinical information or focused entirely on hospitalised patients, limiting extrapolation to other clinical settings.

Secondly, We had a systematic approach to testing our patient cohort recruiting both proven and strongly suspected cases in a consecutive manner, limiting bias. We also collected detailed clinical metadata on our patient cohort, allowing us to look for patient level differences in antibody response. Finally, anonymised data including the index cut-off is available from our paper, to rapidly allow meta-analysis with other cohorts.

One weakness of our paper is our limited data on healthcare worker clinical information. Due to data protection concerns, we did not have access to individual data, although to our knowledge very few healthcare workers locally had severe disease, so they are likely to represent a cohort of milder disease. To our knowledge, less than five healthcare workers were admitted during the study period, suggesting this cohort can be extrapolated to those with milder disease.

### Implications

This paper suggests the sensitivity of the Abbott Architect IgG assay, a PHE approved assay, is significantly worse than previously reported in the cohort it is most likely to be useful in: individuals with mild disease. Clinicians should not rely on a negative test ruling out exposure to SARS-CoV-2 in this setting. Further research should rapidly identify if this is a failure of patients with mild disease to produce significant IgG, as suggested by some, or simply an insensitive assay. Finally, further research should identify the role of antibody testing in suspected PCR negative cases, and the differential performance of PCR diagnostic systems.

## Conclusion

The Abbot Architect SARS-CoV-2 assay has significantly variable performance depending on the clinical cohort. The sensitivity of the assay in individuals with milder disease is lower than in hospitalized patients. Further studies should compare this with other platforms, and assess the clinical utility of this test.

## Data Availability

Data including time from symptom onset, patient status, and index cut off are available from the authors GitHub: https://github.com/gushamilton/discover_serology

## Data sharing

Data including time from symptom onset, patient status, and index cut off are available from the author’s GitHub: https://github.com/gushamilton/discover_serology

## Ethics

Ethical approval for DISCOVER was approved by the HRA: REC (20/YH/0121). Ethical approval for the Pleural Investigation Database was approved by the HRA: REC 08/H0102/11. For the staff samples, ethical approval was waived by the hospital trust, as only anonymous data was included.

## Patient and public involvement

Patients and the public were not involved in the design, conduct, or reporting of this study.

## Contributions

FH,DA, APM, PM, MA and NM conceived of the idea. MA, AN, and DW collected clinical samples and performed laboratory governance. PM, BV, and RH performed the assays. EM collected staff data. FH and DA wrote the manuscript, and all authors contributed to reviewing it. FH is the guarantor

